# Dynamic Analysis of Circulating Tumor DNA to Predict the Prognosis and Monitor the Treatment Response of Patients with Metastatic Triple-negative Breast Cancer: a prospective study

**DOI:** 10.1101/2023.07.04.23292206

**Authors:** Yajing Chi, Mu Su, Dongdong Zhou, Fangchao Zheng, Baoxuan Zhang, Ling Qiang, Guohua Ren, Lihua Song, Bing Bu, Shu Fang, Bo Yu, Jinxing Zhou, Jinming Yu, Huihui Li

**Author notes:** Corresponding author: Huihui Li, Department of Breast Medical Oncology, Shandong Cancer Hospital and Institute, Shandong First Medical University and Shandong Academy of Medical Sciences, No.440, Jiyan Road, Huaiyin District, Jinan, Shandong Province, China, Zip: 250017. Jinming Yu, Department of Radiation Oncology, Shandong Cancer Hospital and Institute, Shandong First Medical University and Shandong Academy of Medical Sciences, Jinan, 250017, Shandong Province, China.

## Abstract

**Background:** Limited data are available on the application of circulating tumor DNA (ctDNA) in metastatic triple-negative breast cancer (mTNBC) patients. Here, we investigated the value of ctDNA for predicting the prognosis and monitoring the treatment response in mTNBC patients.

**Methods:** We prospectively enrolled 70 Chinese patients with mTNBC who had progressed after ≤ 2 lines of chemotherapy and collected blood samples to extract ctDNA for 457-gene targeted panel sequencing.

**Results:** Patients with ctDNA+, defined by 12 prognosis-relevant mutated genes, had a shorter progression-free survival (PFS) than ctDNA− patients (5.16 months *vs.* 9.05 months, *P* = 0.001) and ctDNA+ was independently associated with a shorter PFS (HR, 95%CI: 2.67, 1.2–5.96; *P* = 0.016) by multivariable analyses. Patients with a higher mutant-allele tumor heterogeneity (MATH) score (≥ 6.316) or a higher ctDNA fraction (ctDNA% ≥ 0.05) had a significantly shorter PFS than patients with a lower MATH score (5.67 months *vs.*11.27 months, *P* = 0.007) and patients with a lower ctDNA% (5.45 months *vs.* 12.17 months, *P* < 0.001), respectively. Positive correlations with treatment response were observed for MATH score (R = 0.24, *P* = 0.014) and ctDNA% (R = 0.3, *P* = 0.002), but not the CEA, CA125, or CA153. Moreover, patients who remained ctDNA+ during dynamic monitoring tended to have a shorter PFS than those who did not (3.90 months *vs.* 6.10 months, *P* = 0.135).

**Conclusions:** ctDNA profiling provides insight into the mutational landscape of mTNBC and may reliably predict the prognosis and treatment response of mTNBC patients.

**Funding:** This work was supported by the National Natural Science Foundation of China (Grant No. 81902713), Natural Science Foundation of Shandong Province (Grant No. ZR2019LZL018), Breast Disease Research Fund of Shandong Provincial Medical Association (Grant No. YXH2020ZX066), the Start-up Fund of Shandong Cancer Hospital (Grant No. 2020-PYB10), Beijing Science and Technology Innovation Fund (Grant No. KC2021-ZZ-0010-1).

## Introduction

Breast cancer is the most common malignant tumor and the leading cause of cancer-related deaths in women worldwide(Sung et al., 2021). Triple-negative breast cancer (TNBC) represents 15%–20% of all breast cancer cases and exhibits a more aggressive phenotype (with a poorer prognosis) than non-TNBC(Foulkes, Smith, & Reis-Filho, 2010; X. Li et al., 2017; Malorni et al., 2012). Due to the absence of human epidermal growth factor receptor 2 (HER2), estrogen receptor (ER), or progesterone receptor (PR) expression, TNBC lacks effective targeted therapies and treatment regimens. Patients with mTNBC have fewer available treatment options and exhibit worse survival than early-stage TNBC patients. Furthermore, TNBC is a highly heterogeneous disease, resulting in substantial differences in the tumorigenesis, treatment response, and disease progression among patients(Burstein et al., 2015; Jiang et al., 2019; Perou, 2011), which undoubtedly poses great challenges in prognostic prediction of the mTNBC and efficacy assessment for already limited treatment options. Unfortunately, reliable and tailored biomarkers to predict the prognosis and monitor the treatment response of patients with TNBC are yet to be established.

It has been a long-standing clinical management model to predict the prognosis of patients with mTNBC and guide treatment decision-making through imaging examination and mutational features obtained by tumor biopsy. Imaging usually only provides the external characterization of the tumor, but can not reveal tumor internal molecular characteristics. Given the heterogeneity of TNBC, it is not possible to obtain an accurate and comprehensive picture of the mutational landscape using tissue biopsies unless repeated multiple biopsies(Diaz & Bardelli, 2014), and most patients are refractory to repetitive punctures.

Compared with tissue biopsies, “liquid biopsies” collect and analyze tumor-derived substances, such as circulating tumor DNA (ctDNA), circulating tumor cells (CTCs), and exosomes (e.g., from the blood, cerebrospinal fluid, and urine) of cancer patients in a minimally invasive fashion(Alix-Panabières & Pantel, 2016; Palmirotta et al., 2018; Poulet, Massias, & Taly, 2019). It can be used for early diagnosis of tumor patients, predicting tumor recurrence and metastasis, and evaluating the characteristics and clonal evolution of tumor genomes. ctDNA is a specific fraction of cell-free DNA (cfDNA), which is present in the plasma of apoptotic and necrotic tumor cells(Swarup & Rajeswari, 2007). Owing to special biological origin and the potential for multiple repeat sampling, ctDNA is independent of tumor spatial and temporal heterogeneity, convey more valuable information than a conventional tumor biopsy and enable the dynamic monitoring of tumor burden and treatment response(Campos-Carrillo et al., 2020; Chae & Oh, 2019; Dawson et al., 2013; Gerratana et al., 2021). The value of ctDNA in the accurate prediction of drug resistance and clinical outcomes has also been noted(Asante, Calapre, Ziman, Meniawy, & Gray, 2020; Murtaza et al., 2013).

Several studies have demonstrated the prognostic and predictive value of ctDNA for non-mTNBC during or after (neo)adjuvant therapy(Cavallone et al., 2020; H. Kim et al., 2021; Lin et al., 2021; Ortolan et al., 2021; Riva et al., 2017). Previously, researchers have also been relatively circumscribed concentrated on evaluating specific copy number variants (CNVs) or ctDNA-based single mutation or clonal evolution or the ctDNA level to predict the prognosis of mTNBC patients and the efficacy of specific treatment regimens(Barroso-Sousa et al., 2022; Chopra et al., 2020; Collier et al., 2021; Stover et al., 2018; Weber et al., 2021; Wongchenko et al., 2020). Even a study showed that the ctDNA level had no prognostic impact on survival of patients with mTNBC(Madic et al., 2015). A more comprehensive study of the mutational information and related markers embodied in ctDNA as well as a consensus on the predictive role of ctDNA in mTNBC are needed to apply ctDNA in clinical practice.

Hence, in this study, we investigated the mutational characteristics of ctDNA and ctDNA-related markers in mTNBC using targeted, capture-based, next-generation sequencing (NGS), which offers rapid identification and high coverage from a small blood sample. We aimed to dynamically and more comprehensively evaluate the value of ctDNA in predicting the prognosis and monitoring the treatment response of patients with mTNBC.

## Methods

### Study design and sample collection

Between 2018 and 2021, patients with mTNBC who had progressed after ≤ 2 lines of chemotherapy were prospectively enrolled. A 10 mL sample of peripheral blood was collected into an EDTA anticoagulant tube (STRECK Cell-Free DNA BCT^®^) from patients at different time points (i.e., before treatment, during treatment [treatment cycle 3, day 1], and at progression). Within 2 hours of collection, blood samples were centrifuged at 1,600 × g for 10 min at 4 °C to obtain plasma, followed by secondary centrifugation at 16,000 × g for 10 min at 4 °C to obtained peripheral blood cells. Plasma and peripheral blood cells were stored at −80 °C until the extraction of ctDNA and genomic DNA (gDNA). Paraffin-embedded primary or metastatic tumor tissues were collected before treatment and stored at room temperature for later use.

### DNA extraction and targeted capture-based NGS

ctDNA was extracted from peripheral blood using the QIAamp Circulating Nucleic Acid Kit (Qiagen, Germany) while tumor DNA (tDNA) was extracted from the paraffin-embedded tumor tissues using the AllPrep DNA/RNA FFPE Kit (50) (Qiagen, Germany). Normal control gDNA was extracted from white blood cells using the DNeasy Kit (Qiagen, Germany) according to the manufacturer’s instructions. The sequencing library was prepared from the ctDNA and tDNA samples using the KAPA DNA Library Preparation Kit (KAPA Biosystems, USA), while the gDNA sequencing library was constructed using the Illumina TruSeq DNA Library Preparation Kit (Illumina, USA). Library concentration was determined using real-time quantitative PCR and the KAPA Library Quantification Kit (KAPA Biosystems, USA). The library fragments were then size-selected using agarose gel electrophoresis. A targeted NGS panel of 457 genes (**Supplementary Table 1**), which are known to be frequently mutated in tumors, was designed to capture the target DNA fragments. Sequencing libraries were loaded onto a NovaSeq 6000 platform (Illumina, USA) with a 150-bp read length in paired-end mode.

### Sequencing data analysis

Quality control of the raw sequencing data involved the use of FASTP to trim adapters and remove low-quality sequences(Chen, Zhou, Chen, & Gu, 2018). The clean reads were aligned against the Ensemble GRCh37/hg19 reference genome using BWA software(H. Li & Durbin, 2009). PCR duplications were processed using the gencore tool(Chen et al., 2019) and uniquely mapped reads were generated. The SAMtools suite was used to detect single-nucleotide variants (SNVs), insertions, and deletions(H. Li et al., 2009). CNVs were called using CONTRA(J. Li et al., 2012), with copy number ˃ 3 as the threshold of copy number gain and < 1 as the threshold of copy number loss. The maximal tumor somatic variant allelic frequency (max-VAF) describes the highest mutated frequency of ctDNA detected in the cfDNA(Maron et al., 2019). The calculation of the ctDNA fraction (ctDNA%) was based on the autosomal somatic allele fractions. The mutant allele fraction (MAF) and ctDNA% are related as follows: MAF = (ctDNA × 1)/([(1 – ctDNA) × 2] + [ctDNA × 1]); thus, ctDNA = 2/(1/MAF + 1)(Vandekerkhove et al., 2017). The mutant-allele tumor heterogeneity (MATH) score was calculated as the percentage ratio of the width (median absolute deviation [MAD] scaled by a constant factor, so that the expected MAD of a sample from a normal distribution equals the standard deviation) to the center of the distribution of MAFs among the tumor-specific mutated loci; thus, MATH = 100 × MAD/median(Mroz & Rocco, 2013). Tumor mutational burden (TMB) was defined as the number of non-synonymous somatic mutations per megabase of genome examined(Chalmers et al., 2017).

### Statistical analysis

The longest diameter (mm) of the tumor was measured by examining the radiological images. The response evaluation was carried out according to the Response Evaluation Criteria in Solid Tumors (RECIST) guidelines (version 1.1)(Eisenhauer et al., 2009). The Kaplan–Meier method was used for the survival analyses; median comparison was performed using the log-rank test and hazard ratios (HRs) from the Cox proportional hazards model. The optimal cut-off values for ctDNA%, MATH score, and TMB were determined by the R package “survminer”. Progression-free survival (PFS) was defined as the time interval from the initiation of the study to disease progression or death from any cause. Univariate Cox regression was carried out to analyze the mutations related to PFS; only genes mutated in > 5% of the patients were included in the analysis. The correlation between variables was analyzed using the Spearman correlation test and group comparisons were performed using the Wilcoxon rank-sum test. Statistical analysis and data visualization were conducted using R (version 4.0.1). The statistical significance was defined as bilateral *P* < 0.05.

## Results

### Study cohort and sample information

A total of 126 patients with mTNBC were included in the study. Fifty-six patients were excluded, comprising 21 patients who had previously been treated with more than two lines of chemotherapy, 16 patients who were treated with combination therapy (e.g., chemotherapy with radiotherapy), 11 patients who were lost to follow-up, and 8 patients who failed to be evaluated for efficacy. Finally, 139 plasma samples (baseline samples from 70 patients and dynamic samples from 38/70 patients) from 70 patients and paired tumor tissues from 13 patients were collected and sequenced (**Figure 1A, B**). The patient baseline characteristics are shown in **Table 1**. The median age of all patients was 46 (26–75) years. Overall, 82.9% of patients were diagnosed with invasive ductal carcinoma and most of the patients developed visceral metastases at study entry. **Table 2** shows that all patients received chemotherapy-based treatment with the most common chemotherapy drugs, such as gemcitabine, taxane, or platinum. Eleven patients were also treated with immunotherapy. The objective response rate was 38.6%, and 4, 23, 31, and 12 patients had complete response (CR), partial response (PR), stable disease (SD), and progressive disease (PD), respectively. The median PFS (mPFS) for all patients was 6.15 months (**Supplementary Figure 1A**). There was no significant difference in PFS among different treatment lines or regimens (**Supplementary Figure 1B-H**), although patients treated in the first line had a trend towards improved survival compared with those treated in the second and third lines.

**Figure 1.**
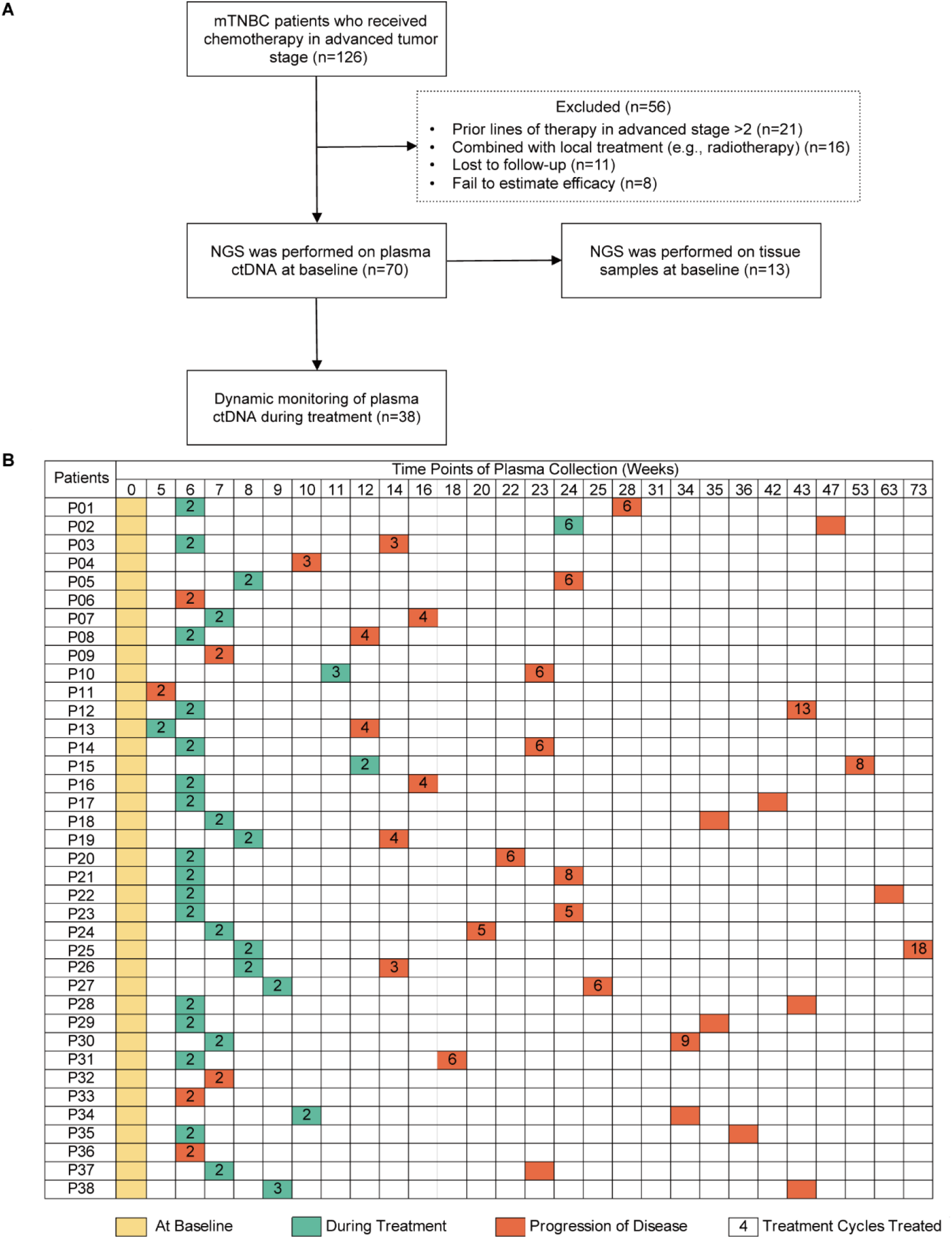
Study design and sample collection. (**A**) Study flowchart. After excluding 82 patients, a total of 70 patients with mTNBC were included in the final analysis. Baseline blood samples were collected from all patients (n = 70) and paired tumor tissues were collected from 13 patients for NGS. (**B**) Blood-sample-derived ctDNA was dynamically monitored at baseline (yellow), during treatment (green), and at progression (orange) for 38 of the 70 patients. ctDNA, circulating tumor DNA; mTNBC, metastatic triple-negative breast cancer; NGS, next-generation sequencing.

**Table 1.** The baseline characteristics in study population.

| Characteristics | All patients-no. (%)<br>(n=70) | Dynamic monitoring patients-no. (%)<br>(n=38) |
| --- | --- | --- |
| **Age (yrs), median (range)** | 46 (26-75) | 47 (27-75) |
| ≤50 | 45 (64.3) | 26 (68.4) |
| >50 | 25 (35.7) | 12 (31.6) |
| **Histopathologic diagnosis** |  |  |
| Invasive ductal carcinoma | 58 (82.9) | 29 (76.3) |
| Other | 10 (14.3) | 7 (18.4) |
| NA | 2 (2.9) | 2 (5.3) |
| **Pathological grade** |  |  |
| I-II | 12 (17.1) | 10 (26.3) |
| III | 39 (55.7) | 20 (52.6) |
| NA | 19 (27.1) | 8 (21.1) |
| **Disease stage at initial diagnosis** |  |  |
| I | 7 (10.0) | 5 (13.2) |
| II | 20 (28.6) | 10 (26.3) |
| III | 33 (47.1) | 20 (52.6) |
| IV | 7 (10.0) | 3 (7.9) |
| NA | 3 (4.3) | 0 |
| **Disease-free interval (months)** |  |  |
| ≤12 (including stage IV at initial diagnosis) | 23 (32.9) | 11 (28.9) |
| >12 | 47 (67.1) | 27 (71.1) |
| **Sites of metastasis** |  |  |
| Visceral | 56 (80.0) | 33 (86.8) |
| Non-visceral | 14 (20.0) | 5 (13.2) |
| **Previous lines of chemotherapy during metastatic stage** |  |  |
| 0 | 46 (65.7) | 22 (57.9) |
| 1 | 19 (27.1) | 11 (28.9) |
| 2 | 5 (7.1) | 5 (13.2) |

**Table 2.** The treatment characteristics and responses of patients.

| Treatment characteristics and responses | All patients-no. (%)<br>(n=70) | Dynamic monitoring patients-no. (%)<br>(n=38) |
| --- | --- | --- |
| **Treatment regimens** |  |  |
| Contained gemcitabine + platinum | 17 (24.3) | 10 (26.3) |
| Contained taxane + platinum | 16 (22.9) | 7 (18.4) |
| Contained vinorelbine + platinum | 4 (5.7) | 1 (2.6) |
| Contained taxane but no platinum | 22 (31.4) | 14 (36.8) |
| Other | 11 (15.7) | 6 (15.8) |
| **Treatment modalities** |  |  |
| Immunotherapy + chemotherapy | 11 (15.7) | 6 (15.8) |
| Chemotherapy | 59 (84.3) | 32 (84.2) |
| **Treatment responses** |  |  |
| CR | 4 (5.7) | 4 (10.5) |
| PR | 23 (32.9) | 9 (23.7) |
| SD | 31 (44.3) | 18 (47.4) |
| PD | 12 (17.1) | 7 (18.4) |

### Mutation characteristics of patients with mTNBC

Plasma samples were obtained from 70 patients with mTNBC before treatment and submitted for targeted NGS. In total, 203 mutated genes were identified using our panel of 457 genes, including 301 missense mutations, 45 frame-shift indels, 16 in-frame indels, 13 splice-site mutations, and 31 stop-gain mutations. The ten most frequently mutated genes were *TP53* (69%), *PIK3CA* (24%), *ARID1A* (9%), *KMT2C* (9%), *CIC* (7%), *KMT2D* (7%), *NOTCH4* (7%), *PBRM1* (7%), *PTEN* (7%), and *DNMT3A* (6%). In addition, gene CNVs were detected in 351/457 genes, 296 of which showed copy number gain (CNG), 38 showed copy number loss, and 17 had both gain and loss mutations. The most prevalent genes with CNVs were *HLA-C* (50%), *HLA-A* (33%), *HLA-B* (33%), *HLA-DRB1* (23%), *SPPL3* (21%), *BTG2* (19%), *C8orf34* (16%), *HLA-DQA1* (16%), *HLA-E* (16%), and *LTBP1* (16%) (**Figure 2A**).

**Figure 2.**
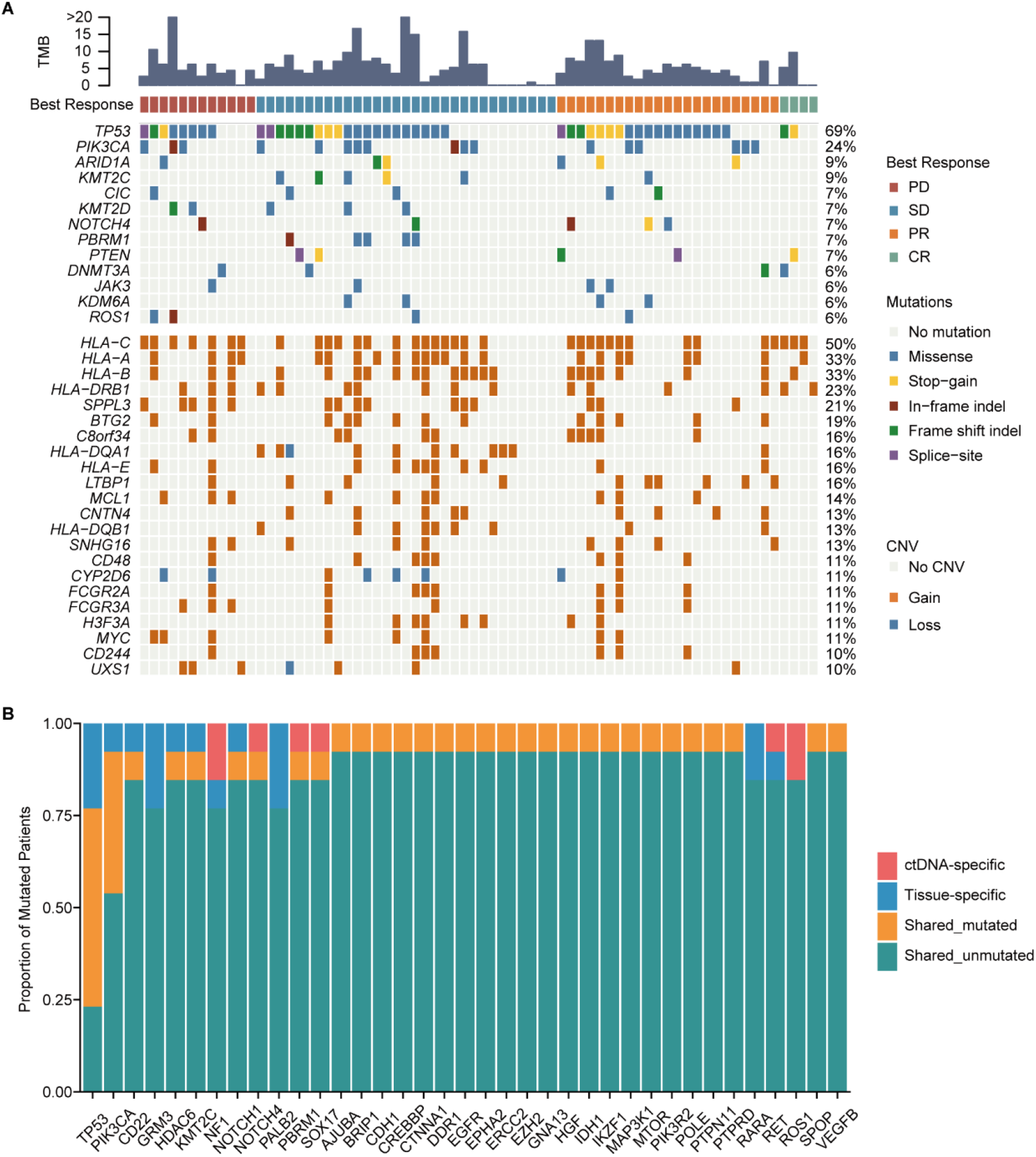
Mutation characteristics of patients with mTNBC. (**A**) The landscape of ctDNA mutations in 70 patients with mTNBC prior to treatment initiation. The patients (n = 70) were divided into four groups (PD, SD, PR, and CR) according to the best treatment response (from left to right). The top half of the figure shows SNVs with a mutation frequency ≥ 5%, and the bottom half shows CNVs with a mutation frequency ≥ 10%; the different colored rectangles represent different types of variation. (**B**) Concordance between the genomic alterations in the blood-derived ctDNA and the tissue-derived tDNA. The mutated genes detected in at least two samples are shown here. The concordance rate = shared mutated genes/(all genes × the number of comparisons) × 100% = (1 – [ctDNA-specific and tissue-specific mutated genes]/[all genes × the number of comparisons]) × 100% = (1 – [37 + 37]/[457 × 13]) × 100% = 98.75%. CNVs, copy number variants; CR, complete response; ctDNA, circulating tumor DNA; tDNA, tumor DNA; mTNBC, metastatic triple-negative breast cancer; PD, progressive disease; PR, partial response; SD, stable disease; SNVs, single-nucleotide variants.

We also used NGS to evaluate the discrepancy and consistency of genomic alterations in ctDNA samples and paired tumor tissues from 13 patients. The mutation frequency in plasma ctDNA was significantly lower than that in the tumor tissues (0.049% ± 0.113% *vs*. 0.168 ± 0.173%, *P* < 0.001) (**Supplementary Figure 2**). A total of 115 mutations in 85 genes were detected, which included 84 mutations in 63 genes from plasma ctDNA and 81 mutations in 55 genes from tDNA. The number of ctDNA-specific and tDNA-specific mutated genes was 37 in both cases. Hence, the concordance rate between mutations in ctDNA and tDNA was 98.75% (**Figure 2B**).

### A ctDNA+/− status correlates with the treatment response and survival of patients with mTNBC

Univariate Cox regression analysis showed that 12 mutated genes, including *HLA-B* (HR, 95% confidence interval [CI]: 1.89, 1.08–3.32), *BTG2* (HR, 95% CI: 2.23, 1.15–4.31), *MCL1* (HR, 95% CI: 2.31, 1.1–4.84), *H3F3A* (HR, 95% CI: 2.36, 1.08–5.16), *MYC* (HR, 95% CI: 3.45, 1.58–7.54), *KMT2C* (HR, 95% CI: 2.75, 1.14–6.63), *KYAT3* (HR, 95% CI: 3.66, 1.48–9.04), *ARID4B* (HR, 95% CI: 4.04, 1.51–10.76), *CD22* (HR, 95% CI: 3.66, 1.25–10.68), *TGFB1* (HR, 95% CI: 3.94, 1.35–11.56), *SGK1* (HR, 95% CI: 3.37, 1.15–9.82), and *RSPO2* (HR, 95% CI: 4.19, 1.42–12.34), indicated a higher risk for recurrence or progression in patients with mTNBC (**Figure 3A**). Moreover, these 12 mutated genes were significantly associated with worse survival (**Supplementary Figure 3**).

**Figure 3.**
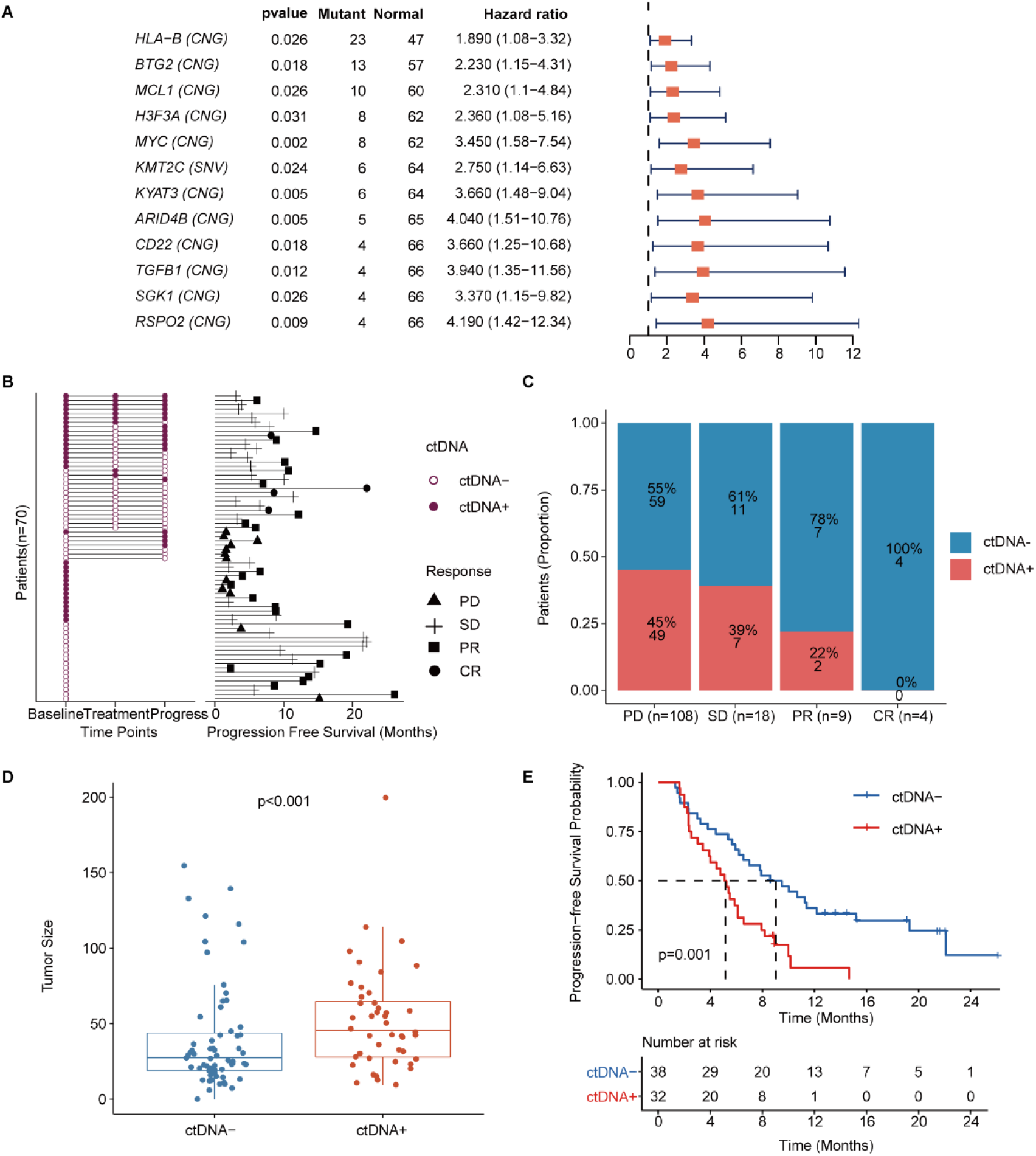
Prognostic relevance of mutations in patients with mTNBC. (**A**) Twelve mutated genes, comprising *HLA-B* (CNG), *BTG2* (CNG), *MCL1* (CNG), *H3F3A* (CNG), *MYC* (CNG), *KMT2C*(SNV), *KYAT3* (CNG), *ARID4B* (CNG), *CD22* (CNG), *TGFB1* (CNG), *SGK1* (CNG), and *RSPO2* (CNG) were identified as being associated with a higher risk of recurrence or progression in patients with mTNBC (all had HRs > 1 and a *P* < 0.05). (**B**) The left half of the figure summarizes the ctDNA status of all patients (n = 70) at different time points; among these, 38 patients also had their ctDNA status recorded during treatment and at progression. Solid dots represent ctDNA+ patients, while unfilled dots represent ctDNA– patients. The length of line segments in the right half of the figure denotes the PFS of patients, whereby the bars indicate the best response (PD, SD, PR, or CR) observed during treatment. (**C**) Comparison of ctDNA status (ctDNA+, red; ctDNA−, blue) among all the blood samples (n = 139) from patients with different treatment responses (PD, SD, PR, or CR). (**D**) The tumor size of the ctDNA+ group at baseline was significantly greater than that of the ctDNA− group at baseline. (**E**) ctDNA+ at baseline was significantly associated with a shorter PFS. CNG, copy number gain; CR, complete response; ctDNA, circulating tumor DNA; ctDNA−, ctDNA negative; ctDNA+, ctDNA positive; HR, hazard ratio; mTNBC, metastatic triple-negative breast cancer; PD, progressive disease; PFS, progression-free survival; PR, partial response; SD, stable disease; SNV, single-nucleotide variant. A *P*-value < 0.05 was used as a measure of statistical significance.

ctDNA was collected and evaluated at different time points and a plasma ctDNA sample with at least one of the 12 prognosis-relevant mutated genes was defined as ctDNA-positive (ctDNA+) (**Figure 3B**). The right half of **Figure 3B** shows that the mPFS in 70 patients with mTNBC was 6.15 months at a median follow-up of 19.13 months. As shown, ctDNA+ patients tended to have a shorter survival duration and less clinical benefit. At baseline, the ctDNA+ rates were 25%, 43%, 55%, and 33% in patients with CR, PR, SD, and PD, respectively. By comparing the ctDNA+/− status at all the time points, we found that the amount of ctDNA+ positively correlated with a worse treatment response. The proportion of ctDNA+ at different time points was 46% (at baseline), 29% (during treatment), and 44% (at progress), while that in the different treatment response groups was 45% (PD), 39% (SD), 22% (PR), and 0% (CR) (**Figure 3C**). Prior to treatment, the tumor size of the ctDNA+ group was significantly larger than that of the ctDNA− group (52.56 ± 34.65 mm *vs*. 40.18 ± 35.49 mm, *P* < 0.001) (**Figure 3D**). We also found that patients who were ctDNA+ at baseline had a shorter PFS than those who were ctDNA− at baseline (5.16 months *vs.* 9.05 months, *P* = 0.001) (**Figure 3E**). Multivariate Cox regression analysis, which included multiple clinical factors and ctDNA status, showed that ctDNA+ was independently associated with a shorter PFS (HR, 95% CI: 2.67, 1.2–5.96; *P* = 0.016) (**Table 3**).

**Table 3.** Multivariate cox regression analysis of multiple clinical factors and ctDNA status with PFS of patients.

| Variable | HR (95 CI) | p value |
| --- | --- | --- |
| Age ($\leq 50$ vs. $> 50$ years) | 1.21 (0.47-3.1) | 0.694 |
| Histopathologic diagnosis (IDC vs. non-IDC) | 0.64 (0.15-2.73) | 0.55 |
| Pathological grade (III vs. I-II) | 1.23 (0.53-2.86) | 0.629 |
| DFI ($\leq 12$months vs. $> 12$ months) | 1.69 (0.6-4.71) | 0.319 |
| **ctDNA status (ctDNA+ vs. ctDNA-)** | **2.67 (1.2-5.96)** | **0.016** |
| T stage (T1 vs. T2 vs. T3) | 0.75 (0.38-1.47) | 0.397 |
| N stage (N1 vs. N2 vs. N3) | 0.79 (0.57-1.1) | 0.161 |
| CEA elevation (Yes vs. No) | 1.75 (0.64-4.81) | 0.278 |
| CA125 elevation (Yes vs. No) | 0.67 (0.24-1.88) | 0.445 |
| CA153 elevation (Yes vs. No) | 1.86 (0.62-5.57) | 0.268 |
| Ki-67 ($\geq 30\%$ vs. $< 30\%$) | 0.78 (0.33-1.88) | 0.582 |
| Site of metastasis (visceral vs. non-visceral) | 0.54 (0.18-1.61) | 0.267 |
| TMB (High vs. Low) | 1.08 (0.38-3.09) | 0.88 |

### Baseline ctDNA-related markers are associated with mTNBC patient survival and treatment response

To further explore the value of ctDNA in predicting clinical outcomes in mTNBC, we examined the association between the pre-treatment ctDNA-related markers (i.e., TMB, MATH score, and ctDNA%) and PFS and the treatment response. Thus, we performed Kaplan–Meier analyses of TMB, MATH score, ctDNA%, and PFS in patients with mTNBC. Although not statistically significant, TMB-high (≥ 2.63) patients tended to have a shorter mPFS than the TMB-low (< 2.63) patients (5.87 months *vs.* 10.03 months, *P* = 0.057) (**Figure 4A**). Meanwhile, patients with a higher MATH score (≥ 6.316) had significantly shorter mPFS than patients with a lower MATH score (< 6.316) (5.67 months *vs.*11.27 months, *P* = 0.007) (**Figure 4B**). Moreover, the higher ctDNA% (≥ 0.05) patient group had a significantly shorter mPFS than the lower ctDNA% (< 0.05) group (5.45 months *vs.* 12.17 months, *P* < 0.001) (**Figure 4C**). Patients with mTNBC were categorized into the PD, SD, PR, and CR groups according to their response to treatment. Further comparative analysis of baseline ctDNA parameters in different treatment response groups revealed that TMB was progressively lower across the four groups, showing a decreasing trend from the PD group to the CR group. Compared with other treatment response groups, the PD group had a larger TMB (*P* = 0.032), greater MATH score (*P* = 0.003), and higher ctDNA% (*P* = 0.002) (**Figure 4D–F**).

**Figure 4.**
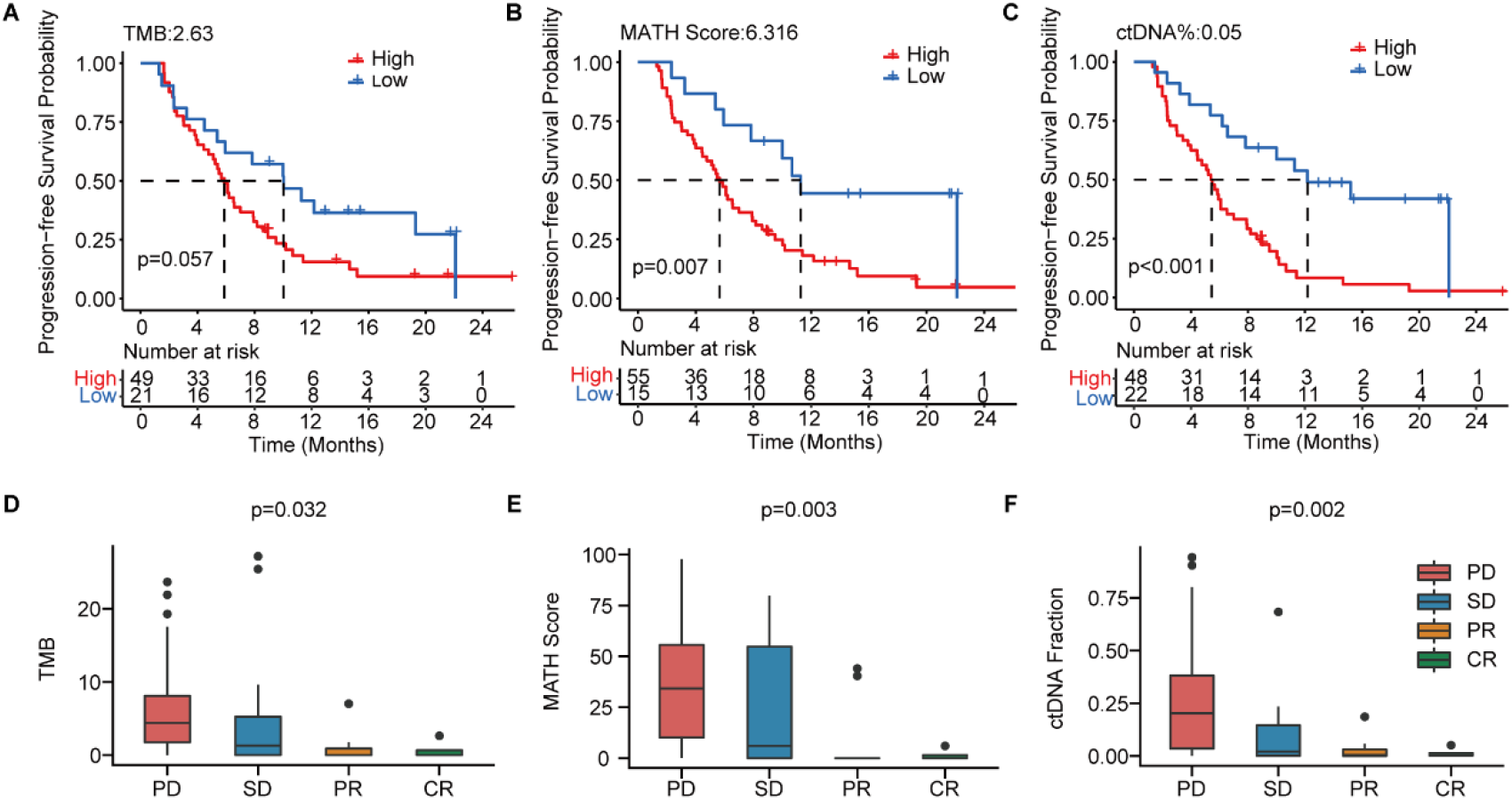
The baseline-ctDNA-derived TMB, MATH score, and ctDNA% were associated with the clinical outcomes of patients with mTNBC. Higher TMB (≥ 2.63) (**A**), MATH score (≥ 6.316) (**B**), and ctDNA% (≥ 0.05) (**C**) were linked to a shorter PFS. The optimal cut-off values for TMB, MATH score, and ctDNA% were determined using the R package “survminer”. Comparison of TMB (**D**), MATH score (**E**), and ctDNA% (**F**) in patients with different treatment responses (PD, SD, PR, or CR). CR, complete response; ctDNA, circulating tumor DNA; ctDNA%, ctDNA fraction; mTNBC, metastatic triple-negative breast cancer; PFS, progression-free survival; PD, progressive disease; PR, partial response; SD, stable disease; TMB, tumor mutational burden. A *P*-value < 0.05 was used as a measure of statistical significance.

### Dynamic changes in ctDNA are associated with treatment response of patients with mTNBC

**Figure 5A–D** highlight the dynamic changes in ctDNA levels (i.e., mutations in 12 prognosis-relevant genes) and traditional tumor markers in each patient with PD (Patient 32), SD (Patient 31), PR (Patient 29), or CR (Patient 18). For instance in Patient 32, the MAF of *MYC* ctDNA increased significantly and was accompanied by increased CA125 and CA153 levels and decreased CEA levels at the time of disease progression (**Figure 5A**). **Figure 5B–D** shows evidence of ctDNA mutations in Patient 31 (*BTG2*, *ARID4B*, *CD22*, *H3F3A*, *HLA-B*, *MCL1*, *MYC*, *RSPO2*), Patient 29 (*BTG2*, *ARID4B*, *H3F3A*, *HLA-B*, *MCL1*, *MYC*), and Patient 18 (*BTG2*, *ARID4B*, *H3F3A*, *HLA-B*, *SGK1*); their mutational rates dropped to the lowest level during the best response to treatment and rose again during progression. CA125 levels varied in line with treatment response and progression but no similar fluctuations were observed for CA153 and CEA. However, compared with the traditional tumor markers, dynamic changes in ctDNA mutations seemed to better mirror treatment-induced changes in tumor size. We therefore analyzed the correlation between the levels of serum tumor markers and tumor size on computed tomography (CT) scans during treatment (**Figure 5E**). We found that tumor size positively correlated with the MATH score (R = 0.24, *P* = 0.014) and ctDNA% (R = 0.3, *P* = 0.002), but not CEA, CA125, or CA153 levels. There were also strong positive correlations among the three ctDNA-related markers, TMB, MATH score, and ctDNA%. Moreover, the dynamic changes in ctDNA status may predict the prognosis of mTNBC. Kaplan–Meier analysis found that patients who remained ctDNA+ during dynamic monitoring had a shorter PFS than those who did not (3.90 months *vs.* 6.10 months, *P* = 0.135) (**Figure 5F**); however, this difference did not achieve statistical significance, most likely due to the limited sample size.

**Figure 5.**
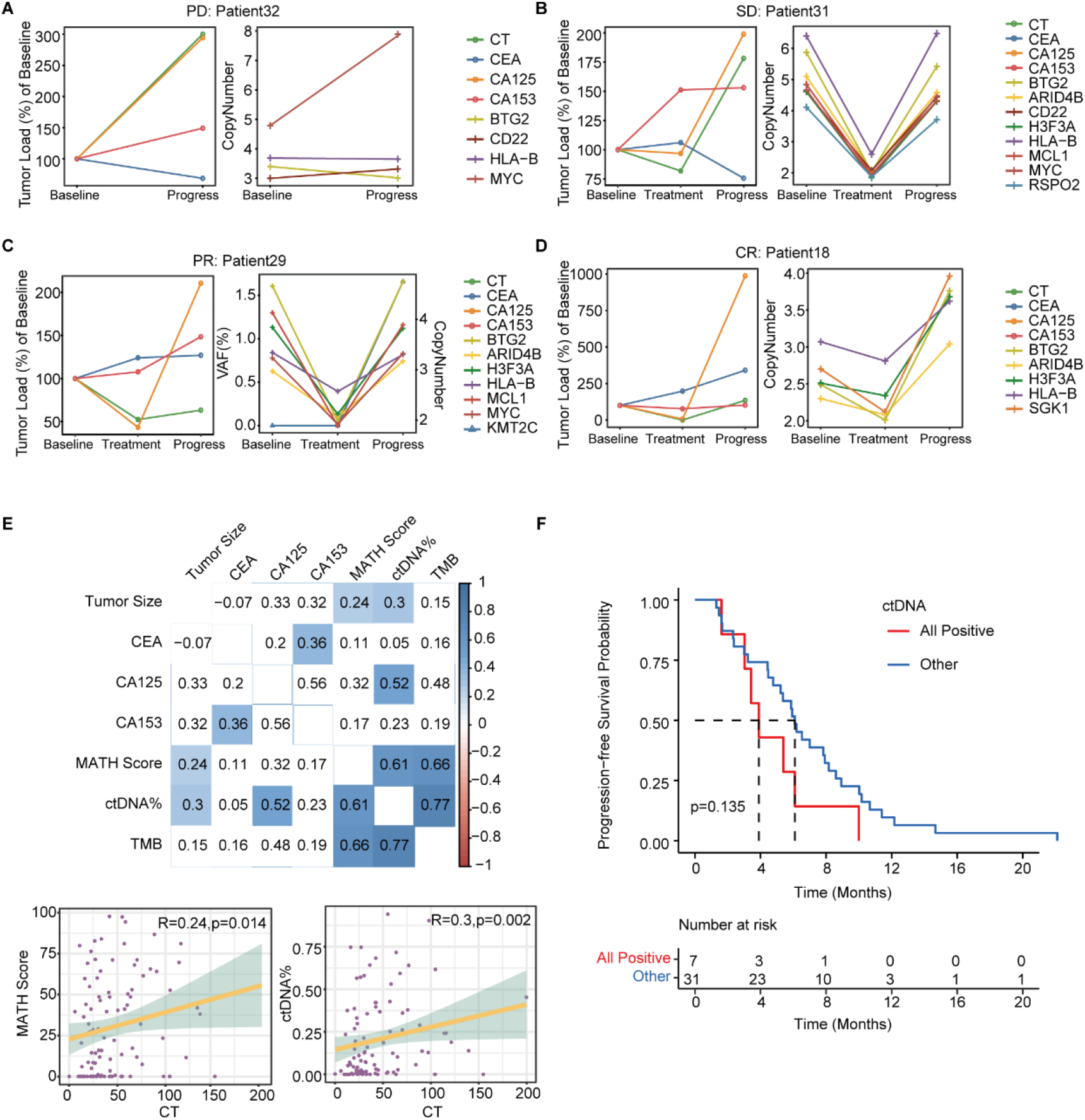
Dynamic ctDNA changes in patients with mTNBC. (**A–D**) Dynamic changes in tumor size were observed using computed tomography (CT) scans and the conventional tumor markers CEA, CA125, and CA153 (left side) or the VAF/copy number of 12 prognosis-relevant genes (right side) in four patients who each had a different best treatment response: PD (Patient 32), SD (Patient 31), PR (Patient 29), and CR (Patient 18). (**E**) The correlation between tumor size (measured using CT scans) and conventional tumor markers (CEA, CA125, CA153) or ctDNA parameters (TMB, MATH score, ctDNA%). Blue: positive correlation; red: negative correlation (the stronger the correlation, the darker the color). (**F**) Patients with a ctDNA+ status across all time points (All positive) tended to have a shorter PFS than those who were ctDNA− at least once during the process of dynamic monitoring (Other). CR, complete response; ctDNA, circulating tumor DNA; ctDNA−, ctDNA negative; ctDNA+, ctDNA positive; ctDNA%, ctDNA fraction; mTNBC, metastatic triple-negative breast cancer; PFS, progression-free survival; PD, progressive disease; PR, partial response; SD, stable disease; TMB, tumor mutational burden; VAF, variant allelic frequency. A *P*-value < 0.05 was used as a measure of statistical significance.

## Discussion

Since the advent of precision medicine, liquid biopsies have become more widely utilized in the clinical management of cancer. In recent years, ctDNA has become the focus of extensive research as a predictive biomarker of tumor progression and/or treatment response. Several studies have explored the promising applications of ctDNA in breast cancer. For example, some researchers have investigated the longitudinal dynamics of ctDNA in the treatment monitoring of metastatic breast cancer, while others have studied the prognostic and predictive value of ctDNA during neoadjuvant chemotherapy for TNBC(Cavallone et al., 2020; Gerratana et al., 2021; Ortolan et al., 2021; Riva et al., 2017). However, the application of ctDNA in monitoring mTNBC is rare in clinical practice.

In the current study, we performed targeted, capture-based NGS (with a 457-gene panel) on 139 plasma samples obtained by liquid biopsy from 70 patients with mTNBC. Thirteen paired tumor tissues were also analyzed to verify if ctDNA could be a feasible alternative to tumor-tissue-derived tDNA. This study demonstrated how a ctDNA-based platform could reliably reveal mutational profiles, stably predict the prognosis, and consistently monitor the treatment response of patients with mTNBC. This study also resolved the uncertainty of some current studies regarding the value of ctDNA and provided clear ctDNA-related predictive markers for mTNBC patients. By evaluating the mutational landscape of mTNBC in the Chinese population using ctDNA analysis, we showed that the most frequently mutated genes were *TP53*, *PIK3CA*, *ARID1A*, and *KMT2C*, and the most frequent CNV was detected in *HLA-C*, *HLA-A*, *HLA-B*, and *HLA-DRB1* (**Figure 2A**). These genes play significant roles in the tumorigenesis, progression, invasion, and metastasis of breast cancer. In the homogeneous population, the most common mutations detected in tumor tissue were *TP53*, followed by *PIK3CA*, *KMT2C*, and *PTEN*; however, the top-ranking CNVs were found in different genes to those identified in a previous report: *E2F3*, *IRS2*, *CCNE1*, *EGFR*, *NFIB*, *CCND1*, and *MYB*(Jiang et al., 2019). This difference may be due to the fact that the plasma-derived ctDNA contains smaller fragments than those that are typically found in tissue-derived tDNA. Meanwhile, compared with the results of other ctDNA identification studies, the distribution of frequent variants in the TNBC cohort was overall consistent(Davis et al., 2020; Rong et al., 2020; Wang et al., 2021). In the study by Chae et al., the concordance between all ctDNA- and tDNA-derived genes was also similar to that in our study (91.0%–94.2% *vs*. 98.75%)(Chae et al., 2017) (**Figure 2B**). The authors reported that the ctDNA-based assay had a high specificity, with a diagnostic accuracy of up to 80%. Although the mutation frequency of tDNA was higher than that of ctDNA in our study, the number of mutations detected in both types of DNA was similar. In addition, we detected 37 specific mutations in each of the ctDNA and tDNA groups, demonstrating the complementarity of blood-derived ctDNA and tissue-derived tDNA. Moreover, compared with tumor tissue analysis, ctDNA assays only require a small blood sample, can capture a variety of mutations (including SNVs and CNVs), and provide information on spatial tumor heterogeneity.

Using NGS, we identified 12 prognosis-relevant mutated genes, which were associated with the shorter PFS of patients with mTNBC (**Supplementary Figure 3**). Most of the 12 genes have been linked to breast cancer by previous studies. For instance, the aberrant expression of *KMT2C* (low expression) and *ARID4B* (high expression) contribute to the poor prognosis of patients with ER-positive breast cancer(Sato & Akimoto, 2017; J. Zhang et al., 2021). However, previous studies have shown that the upregulated expression of *BTG2* and *CD22* were associated with improved survival, which is not in agreement with our current findings(Mascia et al., 2022; Y. J. Zhang et al., 2013); this may be due to the low number of patients included in this study. *TGFB1*, *SGK1*, *RSPO2,* and *MCL1* are implicated in the invasion, migration, growth, autophagy, and progression of TNBC, while *RSPO2* and *MCL1* overexpression is associated with shorter survival rates in patients with TNBC(Coussy et al., 2017; S. Kim, Lee, Jeon, Nam, & Lee, 2015; Yang et al., 2014; Zhu et al., 2020).

As a common driver of breast cancer, *MYC* amplification plays a role in emerging or acquired chemotherapy resistance during neoadjuvant treatment of TNBC and can also synergize with *MCL1* amplification to maintain chemoresistance(Lee et al., 2017). *HLA-B*, a major histocompatibility complex (MHC) class I molecule, is involved in immunosurveillance against tumors and its expression is correlated with the invasiveness and prognosis of breast cancer(Concha, Esteban, Cabrera, Ruiz-Cabello, & Garrido, 1991). To date, there have been no reports of an association between *H3F3A* or *KYAT3* and breast cancer. Previously, *H3F3A* was identified as a driver gene in glioma, and its overexpression is linked to shorter survival rates and disease progression in lung cancer(Felker & Broniscer, 2020; Park et al., 2016). Thus, although the associations between of some of the 12 mutated genes identified in our study and cancer are known, their roles in the prognosis of mTNBC need to be further defined.

We also explored the value of ctDNA status as a biomarker for predicting the prognosis and monitoring the treatment response of patients with mTNBC. We found that a ctDNA+ status was associated with a worse treatment response (**Figure 3B, C**). In addition, we showed that the ctDNA status at baseline could potentially discriminate between mTNBC patients with a high or low lesion load (**Figure 3D**), predict their prognosis (**Figure 3E**), and act as an independent prognostic factor (**Table 3**). This indicates that the ctDNA status, associated with the presence of the 12 prognosis-relevant mutated genes, may be a good guide to the prediction of clinical outcomes and the clinical management of TNBC. We further explored the optimal cut-off values for the ctDNA-based TMB, MATH score, and ctDNA% parameters at baseline using Kaplan–Meier analysis. A high MATH score (≥ 6.316) and high ctDNA% (≥ 0.05) were associated with a significantly shorter PFS (**Figure 4B, C**) and may therefore be related to the tumor burden of mTNBC. A study reported that most ctDNA fragments originate from metastases and not early-stage cancer, suggesting that ctDNA-based NGS may be more suitable for the analysis of metastatic tumors(Vandekerkhove et al., 2017). Unlike ctDNA%, the MATH score represents tumor heterogeneity. A previous study found that TNBC was associated with a higher MATH score(Ma, Jiang, Liu, Liu, & Shao, 2017). Moreover, patients with higher MATH (“Clinical practice guidelines for the use of tumor markers in breast and colorectal cancer. Adopted on May 17, 1996 by the American Society of Clinical Oncology,” 1996)scores tend to have more diverse tumor cell clones and may be more prone to drug resistance and progression(McDonald et al., 2019; Mroz & Rocco, 2013). Thus, the MATH score could also potentially be used as a biomarker for mTNBC prognosis.

Breast cancer is a highly heterogeneous and dynamic disease; therefore, longitudinal monitoring and management are necessary(Garcia-Murillas et al., 2015). The predictive value of ctDNA has prompted further exploration of its feasibility in the dynamic monitoring of the efficacy of neoadjuvant therapy for breast cancer, and in predicting the occurrence of distal metastasis and drug resistance(Cavallone et al., 2020; Darrigues et al., 2021; Wang et al., 2021). Here, we monitored ctDNA to track the dynamic changes in the 12 identified prognosis-related genes during the treatment of patients with mTNBC. The results showed that the elimination of these mutations or the reduction in the mutation rate of these genes was often associated with a better treatment response. Conversely, reappearance of these mutations at a later time point or an increase in their mutation rate signaled disease progression. Thus, we showed that the analysis of ctDNA was sensitive and accurately reflected the treatment response and disease status of patients with mTNBC in a timely manner. Conventional tumor markers have been widely used in clinical cancer management for some time(“Clinical practice guidelines for the use of tumor markers in breast and colorectal cancer. Adopted on May 17, 1996 by the American Society of Clinical Oncology,” 1996). We found that, the serum CEA and CA153 levels contradicted the treatment response of Patient 31 at the mid-treatment time point (**Figure 5B**). The elevation of conventional tumor markers could indicate changes in the tumor and could be interpreted as an early warning of disease progression or pseudo-progression. However, this pseudo-progression or “tumor marker spike” is actually caused by extensive neoplastic cell necrosis induced by anti-tumor therapy. This phenomenon is observed in up to 30% of patients who respond to treatment(Seregni, Coli, & Mazzucca, 2004). NGS-mediated ctDNA detection identifies hundreds or thousands of mutations. Even if individual mutations were the result of a “ctDNA spike” (similar to a “tumor marker spike”), other mutations could be relied upon to accurately signal treatment efficacy. Moreover, we observed positive correlations between treatment response and MATH score and ctDNA%, but not the CEA, CA125, and CA153 levels (**Figure 5E**). Hence, compared with conventional tumor markers, ctDNA dynamics may better reflect treatment response and progression in mTNBC.

Several limitations exist in our study. First, this was a single-center study with a small sample size. Second, the relatively short median follow-up duration was insufficient for capturing a clinically significant association between ctDNA mutations and overall survival. Third, several patients were lost to follow-up, which may have biased the results. Fourth, the potential influence of different treatment lines and regimens was not evaluated; nevertheless, no differences were found in survival based on these factors. Finally, compared with whole-exome sequencing or whole-genome sequencing, NGS with a panel of 457 selected genes, as used in our study, provided limited mutation data.

## Conclusions

ctDNA profiling is a good alternative to tumor tissue sequencing and provides valuable insights into the mutational landscape of mTNBC. Furthermore, our study addressed the value of ctDNA in predicting the prognosis and monitoring the treatment response of patients with mTNBC. The results revealed that higher ctDNA%, MATH score, TMB, ctDNA+ status, and mutation rate were associated with a poor prognosis and a worse treatment response in mTNBC. Moreover, the longitudinal monitoring of genetic biomarkers in ctDNA was more sensitive and accurate for discerning treatment response or progression than traditional tumor markers such as CEA, CA125, and CA153. Taken together, these findings will contribute to a better understanding of ctDNA in mTNBC and may facilitate the development of a more accurate and non-invasive clinical strategy for managing patients with this condition. However, larger clinical trials are necessary to validate our results.

## Additional information

### Funding

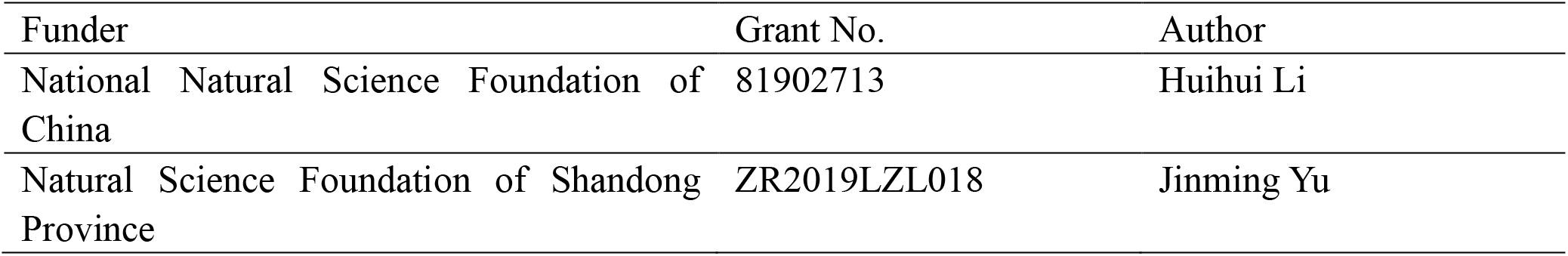
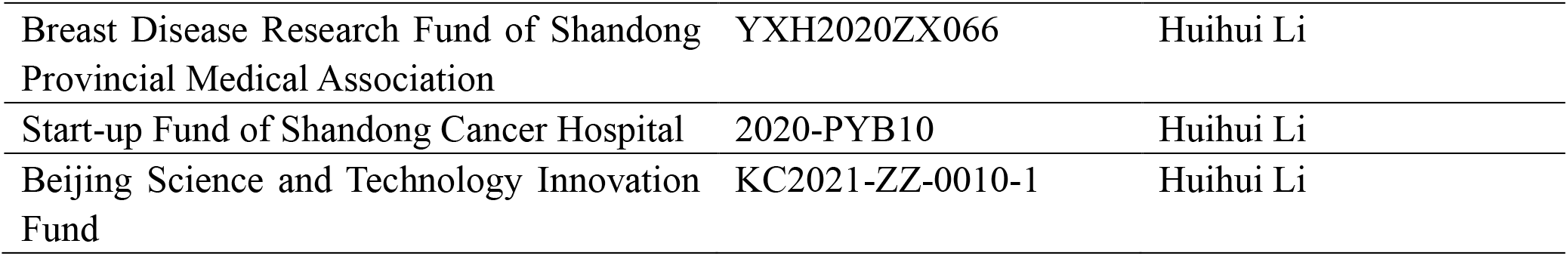

## Data Availability

The raw sequence data generated during the current study have been deposited in the China National Genomics Data Center (https://ngdc.cncb.ac.cn/gsa-human/). The data under accession HRA002598 will be available on 27 June 2024 and are also available from the corresponding author upon reasonable request.

## Acknowledgments

We sincerely thank the support of Yongsheng Wang, Pengfei Qiu, Binbin Cong, Peng Chen, Yanbing Liu, Chunjian Wang, Zhaopeng Zhang, Tong Zhao, Xiao Sun, Zhiyong Yu, Zhijun Huo, Xinzhao Wang, Shubin Song, Liang Zhang, Zhaoyun Liu, Fukai Wang, Chao Li, Xiang Song, Wenshu Zuo, Hui Fu, Meizhu Zheng, Ben Yang, Chao Han, Qian Shao, Xijun Liu, Jinzhi Wang, Wei Wang, Fengxiang Li, Yun Zhao, Linlin Wang, Bingjie Fan, Bing Zou, Zhenhua Gao, Xiangjiao Meng, Liyang Jiang, Zhengqiang Yang and Peng Xie. We also thank Liwen Bianji (Edanz) (www.liwenbianji.cn) for editing a draft of this manuscript.

## Ethics

Human subjects: The study was approved by the Ethics Committee of Shandong Cancer Hospital and Institute (approval number: SDTHEC201806003) that collection of information, tumor tissues and blood samples within the ethical limitition of the patients, and conducted according to the Declaration of Helsinki. Written informed consent was obtained from all patients.

---

**Supplement table 1.**
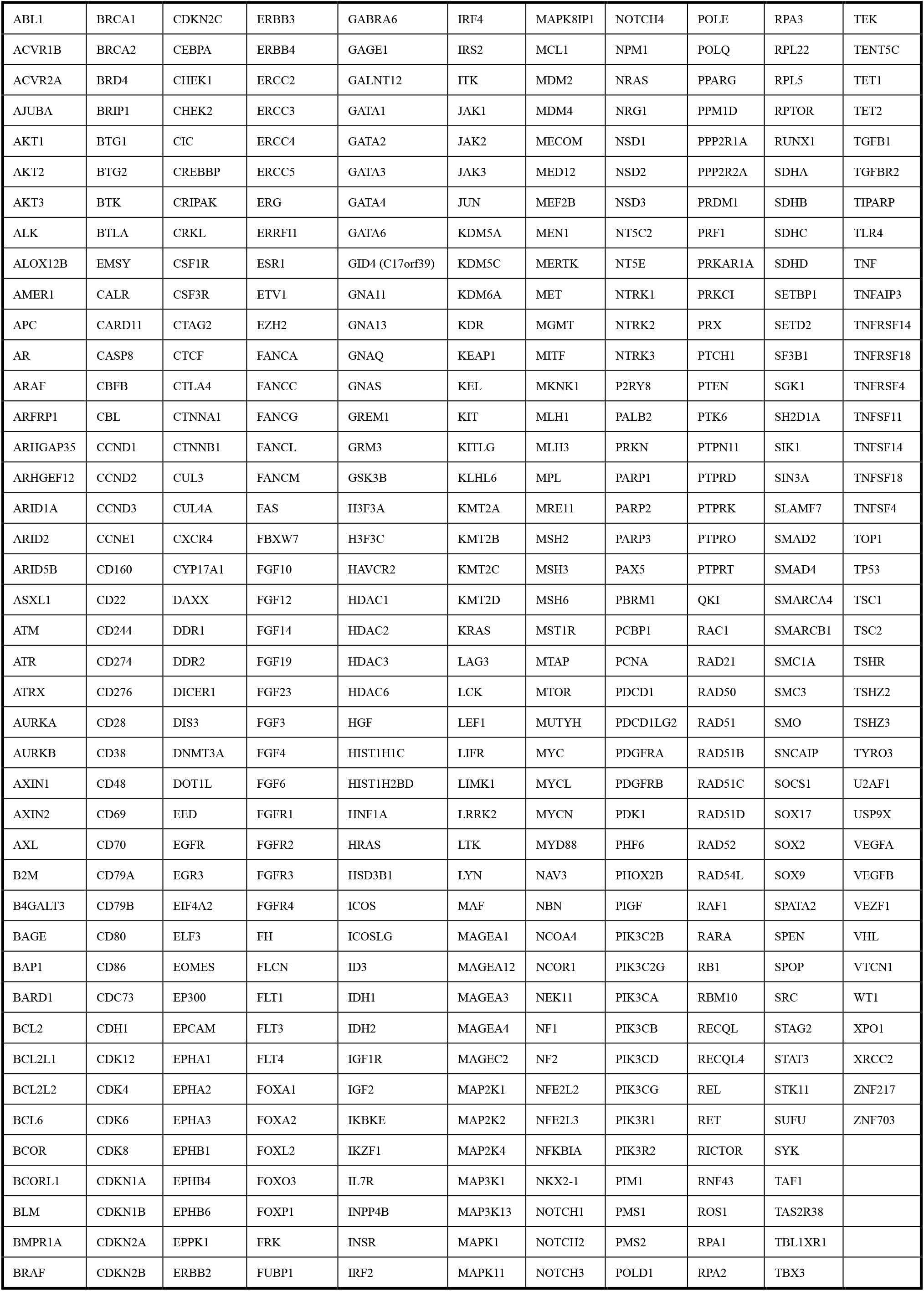
The list of 457 genes detected in this study.

**Supplementary Figure 1.**
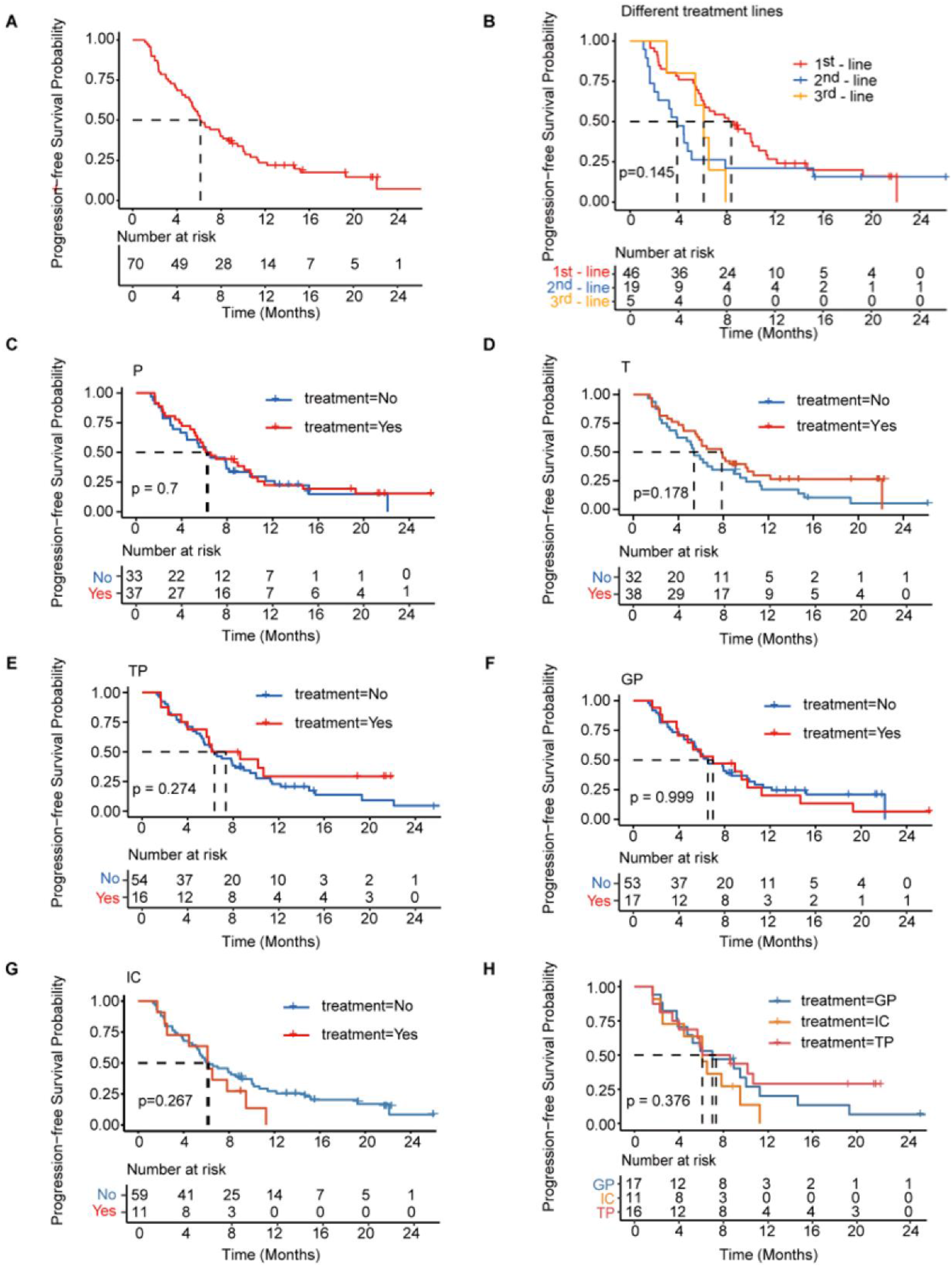
(**A**) Kaplan–Meier analysis of PFS in all patients with mTNBC. (**B**) Kaplan–Meier analysis of PFS in patients with different lines of treatment. (**C–H**) Kaplan–Meier analysis of PFS in patients treated with distinct treatment regimens. GP, gemcitabine- and platinum-based treatment; IC, immunotherapy plus chemotherapy; mTNBC, metastatic triple-negative breast cancer; P, platinum-based treatment; PFS, progression-free survival; T, taxane-based treatment; TP, taxane- and platinum-based treatment. A *P*-value < 0.05 was used as a measure of statistical significance.

**Supplementary Figure 2.**
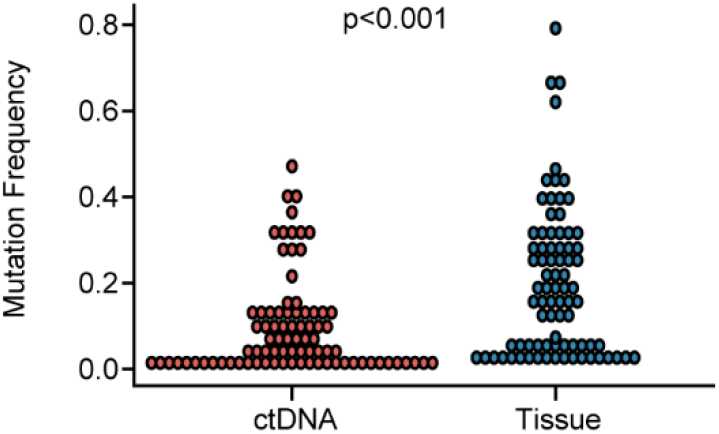
Comparison of mutation frequency between ctDNA and tissue. The mutation frequency in ctDNA was significantly lower than that in tissues (*P* < 0.001). ctDNA, circulating tumor DNA. A *P*-value < 0.05 was used as a measure of statistical significance.

**Supplementary Figure 3.**
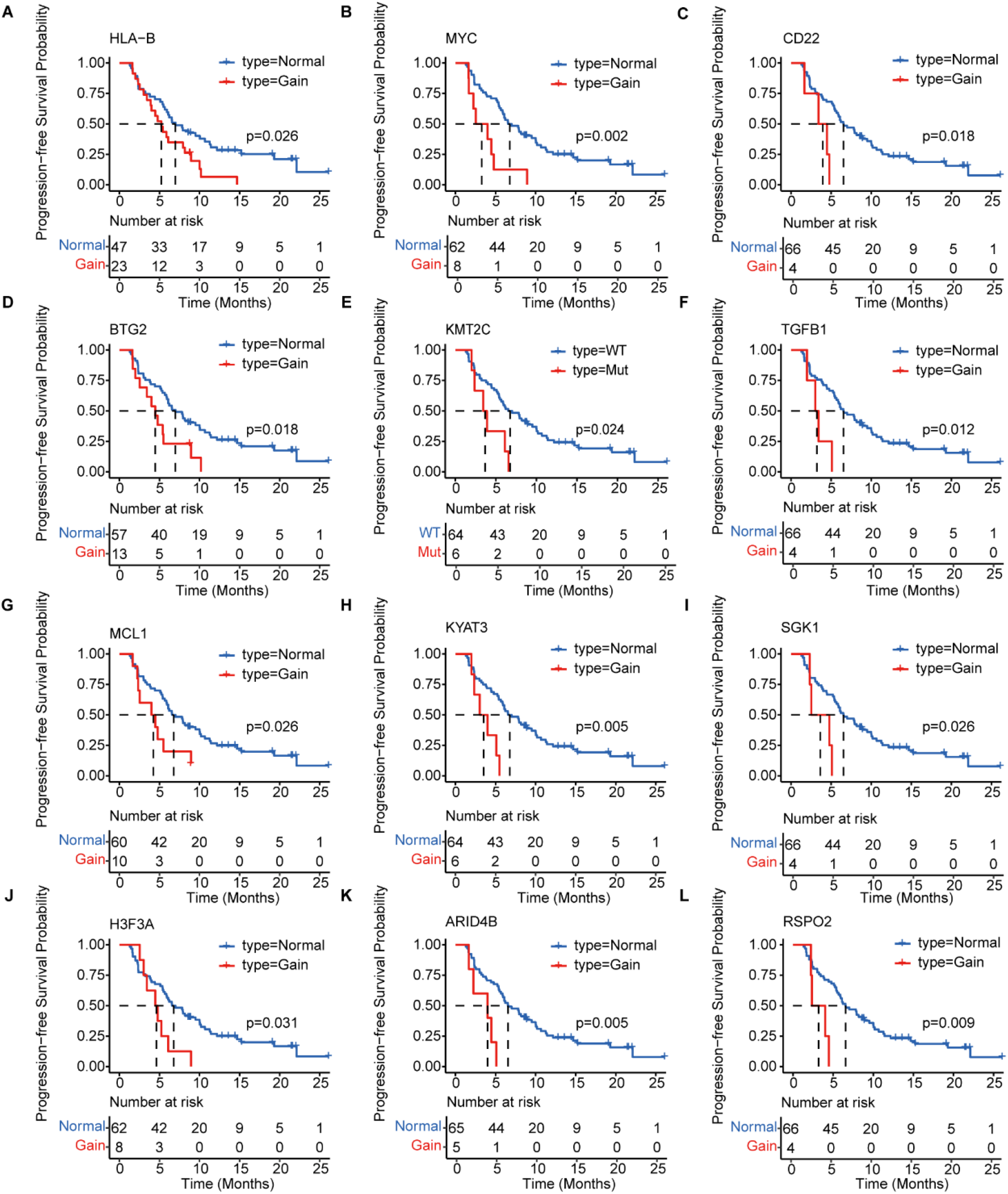
Kaplan–Meier analysis of the PFS of 12 prognosis-relevant mutated genes in patients with mTNBC. Patients with mutated *HLA-B* (**A**), *MYC* (**B**), *CD22* (**C**), *BTG2* (**D**), *KMT2C* (**E**), *TGFB1* (**F**), *MCL1* (**G**), *KYAT3* (**H**), *SGK1* (**I**), *H3F3A* (**J**), *ARID4B* (**K**), or *RSPO2* (**L**) had a significantly shorter PFS. Gain = copy number gain; mTNBC, metastatic triple-negative breast cancer patients; Mut, mutated; PFS, progression-free survival. WT, wild type. A *P*-value < 0.05 was used as a measure of statistical significance.

## References

1. Alix-Panabières, C., & Pantel, K. (2016). Clinical Applications of Circulating Tumor Cells and Circulating Tumor DNA as Liquid Biopsy. Cancer Discov, 6(5), 479–491. doi:10.1158/2159-8290.Cd-15-1483

2. Asante, D. B., Calapre, L., Ziman, M., Meniawy, T. M., & Gray, E. S. (2020). Liquid biopsy in ovarian cancer using circulating tumor DNA and cells: Ready for prime time? Cancer Lett, 468, 59–71. doi:10.1016/j.canlet.2019.10.014

3. Barroso-Sousa, R., Forman, J., Collier, K., Weber, Z. T., Jammihal, T. R., Kao, K. Z., … Tolaney, S. M. (2022). Multidimensional Molecular Profiling of Metastatic Triple-Negative Breast Cancer and Immune Checkpoint Inhibitor Benefit. JCO Precis Oncol, 6, e2100413. doi:10.1200/po.21.00413

4. Burstein, M. D., Tsimelzon, A., Poage, G. M., Covington, K. R., Contreras, A., Fuqua, S. A., … Brown, P. H. (2015). Comprehensive genomic analysis identifies novel subtypes and targets of triple-negative breast cancer. Clin Cancer Res, 21(7), 1688–1698. doi:10.1158/1078-0432.Ccr-14-0432

5. Campos-Carrillo, A., Weitzel, J. N., Sahoo, P., Rockne, R., Mokhnatkin, J. V., Murtaza, M., … Slavin, T. P. (2020). Circulating tumor DNA as an early cancer detection tool. Pharmacol Ther, 207, 107458. doi:10.1016/j.pharmthera.2019.107458

6. Cavallone, L., Aguilar-Mahecha, A., Lafleur, J., Brousse, S., Aldamry, M., Roseshter, T., … Basik, M. (2020). Prognostic and predictive value of circulating tumor DNA during neoadjuvant chemotherapy for triple negative breast cancer. Sci Rep, 10(1), 14704. doi:10.1038/s41598-020-71236-y

7. Chae, Y. K., Davis, A. A., Jain, S., Santa-Maria, C., Flaum, L., Beaubier, N., … Cristofanilli, M. (2017). Concordance of Genomic Alterations by Next-Generation Sequencing in Tumor Tissue versus Circulating Tumor DNA in Breast Cancer. Mol Cancer Ther, 16(7), 1412–1420. doi:10.1158/1535-7163.Mct-17-0061

8. Chae, Y. K., & Oh, M. S. (2019). Detection of Minimal Residual Disease Using ctDNA in Lung Cancer: Current Evidence and Future Directions. J Thorac Oncol, 14(1), 16–24. doi:10.1016/j.jtho.2018.09.022

9. Chalmers, Z. R., Connelly, C. F., Fabrizio, D., Gay, L., Ali, S. M., Ennis, R., … Frampton, G. M. (2017). Analysis of 100,000 human cancer genomes reveals the landscape of tumor mutational burden. Genome Med, 9(1), 34. doi:10.1186/s13073-017-0424-2

10. Chen, S., Zhou, Y., Chen, Y., & Gu, J. (2018). fastp: an ultra-fast all-in-one FASTQ preprocessor. Bioinformatics, 34(17), i884–i890. doi:10.1093/bioinformatics/bty560

11. Chen, S., Zhou, Y., Chen, Y., Huang, T., Liao, W., Xu, Y., … Gu, J. (2019). Gencore: an efficient tool to generate consensus reads for error suppressing and duplicate removing of NGS data. BMC Bioinformatics, 20(Suppl 23), 606. doi:10.1186/s12859-019-3280-9

12. Chopra, N., Tovey, H., Pearson, A., Cutts, R., Toms, C., Proszek, P., … Turner, N. C. (2020). Homologous recombination DNA repair deficiency and PARP inhibition activity in primary triple negative breast cancer. Nat Commun, 11(1), 2662. doi:10.1038/s41467-020-16142-7

13. Clinical practice guidelines for the use of tumor markers in breast and colorectal cancer. Adopted on May 17, 1996 by the American Society of Clinical Oncology. (1996). J Clin Oncol, 14(10), 2843–2877. doi:10.1200/jco.1996.14.10.2843

14. Collier, K. A., Asad, S., Tallman, D., Jenison, J., Rajkovic, A., Mardis, E. R., … Stover, D. G. (2021). Association of 17q22 Amplicon Via Cell-Free DNA With Platinum Chemotherapy Response in Metastatic Triple-Negative Breast Cancer. JCO Precis Oncol, 5. doi:10.1200/po.21.00104

15. Concha, A., Esteban, F., Cabrera, T., Ruiz-Cabello, F., & Garrido, F. (1991). Tumor aggressiveness and MHC class I and II antigens in laryngeal and breast cancer. Semin Cancer Biol, 2(1), 47–54.

16. Coussy, F., Lallemand, F., Vacher, S., Schnitzler, A., Chemlali, W., Caly, M., … Bièche, I. (2017). Clinical value of R-spondins in triple-negative and metaplastic breast cancers. Br J Cancer, 116(12), 1595–1603. doi:10.1038/bjc.2017.131

17. Darrigues, L., Pierga, J. Y., Bernard-Tessier, A., Bièche, I., Silveira, A. B., Michel, M., … Bidard, F. C. (2021). Circulating tumor DNA as a dynamic biomarker of response to palbociclib and fulvestrant in metastatic breast cancer patients. Breast Cancer Res, 23(1), 31. doi:10.1186/s13058-021-01411-0

18. Davis, A. A., Jacob, S., Gerratana, L., Shah, A. N., Wehbe, F., Katam, N., … Cristofanilli, M. (2020). Landscape of circulating tumour DNA in metastatic breast cancer. EBioMedicine, 58, 102914. doi:10.1016/j.ebiom.2020.102914

19. Dawson, S. J., Tsui, D. W., Murtaza, M., Biggs, H., Rueda, O. M., Chin, S. F., … Rosenfeld, N. (2013). Analysis of circulating tumor DNA to monitor metastatic breast cancer. N Engl J Med, 368(13), 1199–1209. doi:10.1056/NEJMoa1213261

20. Diaz, L. A., Jr., & Bardelli, A. (2014). Liquid biopsies: genotyping circulating tumor DNA. J Clin Oncol, 32(6), 579–586. doi:10.1200/jco.2012.45.2011

21. Eisenhauer, E. A., Therasse, P., Bogaerts, J., Schwartz, L. H., Sargent, D., Ford, R., … Verweij, J. (2009). New response evaluation criteria in solid tumours: revised RECIST guideline (version 1.1). Eur J Cancer, 45(2), 228–247. doi:10.1016/j.ejca.2008.10.026

22. Felker, J., & Broniscer, A. (2020). Improving long-term survival in diffuse intrinsic pontine glioma. Expert Rev Neurother, 20(7), 647–658. doi:10.1080/14737175.2020.1775584

23. Foulkes, W. D., Smith, I. E., & Reis-Filho, J. S. (2010). Triple-negative breast cancer. N Engl J Med, 363(20), 1938–1948. doi:10.1056/NEJMra1001389

24. Garcia-Murillas, I., Schiavon, G., Weigelt, B., Ng, C., Hrebien, S., Cutts, R. J., … Turner, N. C. (2015). Mutation tracking in circulating tumor DNA predicts relapse in early breast cancer. Sci Transl Med, 7(302), 302ra133. doi:10.1126/scitranslmed.aab0021

25. Gerratana, L., Davis, A. A., Zhang, Q., Basile, D., Rossi, G., Strickland, K., … Cristofanilli, M. (2021). Longitudinal Dynamics of Circulating Tumor Cells and Circulating Tumor DNA for Treatment Monitoring in Metastatic Breast Cancer. JCO Precis Oncol, 5, 943–952. doi:10.1200/po.20.00345

26. Jiang, Y. Z., Ma, D., Suo, C., Shi, J., Xue, M., Hu, X., … Shao, Z. M. (2019). Genomic and Transcriptomic Landscape of Triple-Negative Breast Cancers: Subtypes and Treatment Strategies. Cancer Cell, 35(3), 428–440.e425. doi:10.1016/j.ccell.2019.02.001

27. Kim, H., Kim, Y. J., Park, D., Park, W. Y., Choi, D. H., Park, W., … Kim, N. (2021). Dynamics of circulating tumor DNA during postoperative radiotherapy in patients with residual triple-negative breast cancer following neoadjuvant chemotherapy: a prospective observational study. Breast Cancer Res Treat, 189(1), 167–175. doi:10.1007/s10549-021-06296-3

28. Kim, S., Lee, J., Jeon, M., Nam, S. J., & Lee, J. E. (2015). Elevated TGF-β1 and -β2 expression accelerates the epithelial to mesenchymal transition in triple-negative breast cancer cells. Cytokine, 75(1), 151–158. doi:10.1016/j.cyto.2015.05.020

29. Lee, K. M., Giltnane, J. M., Balko, J. M., Schwarz, L. J., Guerrero-Zotano, A. L., Hutchinson, K. E., … Arteaga, C. L. (2017). MYC and MCL1 Cooperatively Promote Chemotherapy-Resistant Breast Cancer Stem Cells via Regulation of Mitochondrial Oxidative Phosphorylation. Cell Metab, 26(4), 633–647.e637. doi:10.1016/j.cmet.2017.09.009

30. Li, H., & Durbin, R. (2009). Fast and accurate short read alignment with Burrows-Wheeler transform. Bioinformatics, 25(14), 1754–1760. doi:10.1093/bioinformatics/btp324

31. Li, H., Handsaker, B., Wysoker, A., Fennell, T., Ruan, J., Homer, N., Durbin, R. (2009). The Sequence Alignment/Map format and SAMtools. Bioinformatics, 25(16), 2078–2079. doi:10.1093/bioinformatics/btp352

32. Li, J., Lupat, R., Amarasinghe, K. C., Thompson, E. R., Doyle, M. A., Ryland, G. L., … Gorringe, K. L. (2012). CONTRA: copy number analysis for targeted resequencing. Bioinformatics, 28(10), 1307–1313. doi:10.1093/bioinformatics/bts146

33. Li, X., Yang, J., Peng, L., Sahin, A. A., Huo, L., Ward, K. C., Meisel, J. L. (2017). Triple-negative breast cancer has worse overall survival and cause-specific survival than non-triple-negative breast cancer. Breast Cancer Res Treat, 161(2), 279–287. doi:10.1007/s10549-016-4059-6

34. Lin, P. H., Wang, M. Y., Lo, C., Tsai, L. W., Yen, T. C., Huang, T. Y., Huang, C. S. (2021). Circulating Tumor DNA as a Predictive Marker of Recurrence for Patients With Stage II-III Breast Cancer Treated With Neoadjuvant Therapy. Front Oncol, 11, 736769. doi:10.3389/fonc.2021.736769

35. Ma, D., Jiang, Y. Z., Liu, X. Y., Liu, Y. R., & Shao, Z. M. (2017). Clinical and molecular relevance of mutant-allele tumor heterogeneity in breast cancer. Breast Cancer Res Treat, 162(1), 39–48. doi:10.1007/s10549-017-4113-z

36. Madic, J., Kiialainen, A., Bidard, F. C., Birzele, F., Ramey, G., Leroy, Q., … Lebofsky, R. (2015). Circulating tumor DNA and circulating tumor cells in metastatic triple negative breast cancer patients. Int J Cancer, 136(9), 2158–2165. doi:10.1002/ijc.29265

37. Malorni, L., Shetty, P. B., De Angelis, C., Hilsenbeck, S., Rimawi, M. F., Elledge, R., … Arpino, G. (2012). Clinical and biologic features of triple-negative breast cancers in a large cohort of patients with long-term follow-up. Breast Cancer Res Treat, 136(3), 795–804. doi:10.1007/s10549-012-2315-y

38. Maron, S. B., Chase, L. M., Lomnicki, S., Kochanny, S., Moore, K. L., Joshi, S. S., … Catenacci, D. V. T. (2019). Circulating Tumor DNA Sequencing Analysis of Gastroesophageal Adenocarcinoma. Clin Cancer Res, 25(23), 7098–7112. doi:10.1158/1078-0432.Ccr-19-1704

39. Mascia, F., Mazo, I., Alterovitz, W. L., Karagiannis, K., Wu, W. W., Shen, R. F., … Rao, V. A. (2022). In search of autophagy biomarkers in breast cancer: Receptor status and drug agnostic transcriptional changes during autophagy flux in cell lines. PLoS One, 17(1), e0262134. doi:10.1371/journal.pone.0262134

40. McDonald, K. A., Kawaguchi, T., Qi, Q., Peng, X., Asaoka, M., Young, J., … Takabe, K. (2019). Tumor Heterogeneity Correlates with Less Immune Response and Worse Survival in Breast Cancer Patients. Ann Surg Oncol, 26(7), 2191–2199. doi:10.1245/s10434-019-07338-3

41. Mroz, E. A., & Rocco, J. W. (2013). MATH, a novel measure of intratumor genetic heterogeneity, is high in poor-outcome classes of head and neck squamous cell carcinoma. Oral Oncol, 49(3), 211–215. doi:10.1016/j.oraloncology.2012.09.007

42. Murtaza, M., Dawson, S. J., Tsui, D. W., Gale, D., Forshew, T., Piskorz, A. M., … Rosenfeld, N. (2013). Non-invasive analysis of acquired resistance to cancer therapy by sequencing of plasma DNA. Nature, 497(7447), 108–112. doi:10.1038/nature12065

43. Ortolan, E., Appierto, V., Silvestri, M., Miceli, R., Veneroni, S., Folli, S., … Di Cosimo, S. (2021). Blood-based genomics of triple-negative breast cancer progression in patients treated with neoadjuvant chemotherapy. ESMO Open, 6(2), 100086. doi:10.1016/j.esmoop.2021.100086

44. Palmirotta, R., Lovero, D., Cafforio, P., Felici, C., Mannavola, F., Pellè, E., … Silvestris, F. (2018). Liquid biopsy of cancer: a multimodal diagnostic tool in clinical oncology. Ther Adv Med Oncol, 10, 1758835918794630. doi:10.1177/1758835918794630

45. Park, S. M., Choi, E. Y., Bae, M., Kim, S., Park, J. B., Yoo, H., … Kim, I. H. (2016). Histone variant H3F3A promotes lung cancer cell migration through intronic regulation. Nat Commun, 7, 12914. doi:10.1038/ncomms12914

46. Perou, C. M. (2011). Molecular stratification of triple-negative breast cancers. Oncologist, 16 Suppl 1, 61–70. doi:10.1634/theoncologist.2011-S1-61

47. Poulet, G., Massias, J., & Taly, V. (2019). Liquid Biopsy: General Concepts. Acta Cytol, 63(6), 449–455. doi:10.1159/000499337

48. Riva, F., Bidard, F. C., Houy, A., Saliou, A., Madic, J., Rampanou, A., … Pierga, J. Y. (2017). Patient-Specific Circulating Tumor DNA Detection during Neoadjuvant Chemotherapy in Triple-Negative Breast Cancer. Clin Chem, 63(3), 691–699. doi:10.1373/clinchem.2016.262337

49. Rong, G., Yi, Z., Ma, F., Guan, Y., Xu, Y., Li, L., & Xu, B. (2020). Mutational characteristics determined using circulating tumor DNA analysis in triple-negative breast cancer patients with distant metastasis. Cancer Commun (Lond), 40(12), 738–742. doi:10.1002/cac2.12102

50. Sato, K., & Akimoto, K. (2017). Expression Levels of KMT2C and SLC20A1 Identified by Information-theoretical Analysis Are Powerful Prognostic Biomarkers in Estrogen Receptor-positive Breast Cancer. Clin Breast Cancer, 17(3), e135–e142. doi:10.1016/j.clbc.2016.11.005

51. Seregni, E., Coli, A., & Mazzucca, N. (2004). Circulating tumour markers in breast cancer. Eur J Nucl Med Mol Imaging, 31 Suppl 1, S15–22. doi:10.1007/s00259-004-1523-z

52. Stover, D. G., Parsons, H. A., Ha, G., Freeman, S. S., Barry, W. T., Guo, H., … Adalsteinsson, V. A. (2018). Association of Cell-Free DNA Tumor Fraction and Somatic Copy Number Alterations With Survival in Metastatic Triple-Negative Breast Cancer. J Clin Oncol, 36(6), 543–553. doi:10.1200/jco.2017.76.0033

53. Sung, H., Ferlay, J., Siegel, R. L., Laversanne, M., Soerjomataram, I., Jemal, A., & Bray, F. (2021). Global Cancer Statistics 2020: GLOBOCAN Estimates of Incidence and Mortality Worldwide for 36 Cancers in 185 Countries. CA Cancer J Clin, 71(3), 209–249. doi:10.3322/caac.21660

54. Swarup, V., & Rajeswari, M. R. (2007). Circulating (cell-free) nucleic acids--a promising, non-invasive tool for early detection of several human diseases. FEBS Lett, 581(5), 795–799. doi:10.1016/j.febslet.2007.01.051

55. Vandekerkhove, G., Todenhöfer, T., Annala, M., Struss, W. J., Wong, A., Beja, K., … Wyatt, A. W. (2017). Circulating Tumor DNA Reveals Clinically Actionable Somatic Genome of Metastatic Bladder Cancer. Clin Cancer Res, 23(21), 6487–6497. doi:10.1158/1078-0432.Ccr-17-1140

56. Wang, Y., Lin, L., Li, L., Wen, J., Chi, Y., Hao, R., … Wang, O. (2021). Genetic landscape of breast cancer and mutation tracking with circulating tumor DNA in Chinese women. Aging (Albany NY), 13(8), 11860–11876. doi:10.18632/aging.202888

57. Weber, Z. T., Collier, K. A., Tallman, D., Forman, J., Shukla, S., Asad, S., … Stover, D. G. (2021). Modeling clonal structure over narrow time frames via circulating tumor DNA in metastatic breast cancer. Genome Med, 13(1), 89. doi:10.1186/s13073-021-00895-x

58. Wongchenko, M. J., Kim, S. B., Saura, C., Oliveira, M., Lipson, D., Kennedy, M., … Dent, R. (2020). Circulating Tumor DNA and Biomarker Analyses From the LOTUS Randomized Trial of First-Line Ipatasertib and Paclitaxel for Metastatic Triple-Negative Breast Cancer. JCO Precis Oncol, 4, 1012–1024. doi:10.1200/po.19.00396

59. Yang, L., Perez, A. A., Fujie, S., Warden, C., Li, J., Wang, Y., … Yen, Y. (2014). Wnt modulates MCL1 to control cell survival in triple negative breast cancer. BMC Cancer, 14, 124. doi:10.1186/1471-2407-14-124

60. Zhang, J., Hou, S., You, Z., Li, G., Xu, S., Li, X., … Pang, D. (2021). Expression and prognostic values of ARID family members in breast cancer. Aging (Albany NY), 13(4), 5621–5637. doi:10.18632/aging.202489

61. Zhang, Y. J., Wei, L., Liu, M., Li, J., Zheng, Y. Q., Gao, Y., & Li, X. R. (2013). BTG2 inhibits the proliferation, invasion, and apoptosis of MDA-MB-231 triple-negative breast cancer cells. Tumour Biol, 34(3), 1605–1613. doi:10.1007/s13277-013-0691-5

62. Zhu, R., Yang, G., Cao, Z., Shen, K., Zheng, L., Xiao, J., … Zhang, T. (2020). The prospect of serum and glucocorticoid-inducible kinase 1 (SGK1) in cancer therapy: a rising star. Ther Adv Med Oncol, 12, 1758835920940946. doi:10.1177/1758835920940946

